# Deep learning-based algorithm versus physician judgement for diagnosis of myopathy and neuropathy from needle electromyography

**DOI:** 10.1101/2023.01.13.23284511

**Authors:** Ilhan Yoo, Jaesung Yoo, Dongmin Kim, Ina Youn, Hyodong Kim, Michelle Youn, Jun Hee Won, Woosup Cho, Youho Myong, Sehoon Kim, Ri Yu, Sung-Min Kim, Kwangsoo Kim, Seung-Bo Lee, Keewon Kim

## Abstract

Electromyography is a valuable diagnostic procedure for diagnosing patients with neuromuscular diseases; however, it has some drawbacks. First, diagnosis using electromyography is subjective, and in some cases, there is the potential for inter-individual discrepancies. Second, it is a time- and effort-intensive process that requires expertise to yield accurate results. Recently, a deep learning algorithm shows effectiveness for the analysis of waveform data such as electrocardiography. To overcome limitations of electromyography, we developed a deep learning-based electromyography classification system and compared the performance of our deep learning model with that of six physicians. This study included 58 subjects who underwent electromyography and were finally confirmed as having myopathy or neuropathy, or to be in a normal state between June 2015 and July 2020 at Seoul National University Hospital. We developed a one-dimensional convolutional neural network algorithm and divide-and-vote system for diagnosing subjects. Diagnosis results with our deep learning model were compared with those of six physicians with experience in performing and interpreting electromyography. The accuracy, sensitivity, specificity, and positive predictive value of the deep learning model for diagnosis as to whether subjects have myopathy or neuropathy or normal were 0.875, 0.820, 0.904, and 0.820, respectively, whereas those for the physicians were 0.694, 0.537, 0.773, and 0.524, respectively. The area under the receiver operating characteristic curves of the deep learning model for predicting myopathy, neuropathy, and normal states was better than the averaged results of six physicians. Our study showed that deep learning could play a key role in reading electromyography and diagnosing patients with neuromuscular diseases. In the future, large prospective cohort studies incorporating diverse neuromuscular diseases can enable deep learning-based electrodiagnosis on behalf of physicians.

## Introduction

Electromyography (EMG) is an electrophysiological procedure that records electrical activity generated by nerves, muscles, and neuromuscular junctions. EMG is performed via a needle electrode that is inserted into a muscle or surface electrode during resting and volitional states [1–6]. Peripheral nerve and muscle disorders are characterized by EMG signal abnormalities, which reflect the anatomical and physiological states of peripheral nerves and muscles [1–6].

The signal recorded during muscle contraction comprises the motor unit action potential (MUAP), which can be detected from muscles in the volitional state. In a clinical setting, neuropathy and myopathy are difficult to discern because their symptoms can be significantly similar and overlap in some cases, and MUAP plays a significant role in distinguishing them [1,5–12]. Patients with peripheral neuropathy commonly exhibit MUAPs with high amplitudes, long durations, and reduced recruitment, whereas those with myopathy commonly exhibit MUAPs with small amplitudes, short durations, and early recruitment [1,5–12].

Although EMG plays an important role in distinguishing neuromuscular diseases, EMG evaluation has some limitations. First, the accuracy of EMG-based diagnosis is reliant on the examiner’s proficiency. Previous studies have reported an EMG sensitivity of 47–83% for the diagnosis of neuromuscular disease, with a specificity of 73–81% and an interrater reliability of 62–81% [13–15]. Second, considerable time and effort are required to accurately detect EMG signal abnormalities. The increasing prevalence of neuromuscular disease places a burden on physicians owing to the surge in the number of patients requiring EMG [16–19]. An accurate, efficient, and automated approach for reading EMG may help physicians arrive at the diagnosis more readily.

Recently, deep learning has been used to analyze large datasets in several fields. It has been applied to clinical data, including waveform and time-series data, such as electrocardiographic and electroencephalographic data [20–23]. Numerous studies have also investigated the medical applications of deep learning, wherein the performance of the deep learning model was comparable to or surpassed that of humans [24–27]. Previous EMG studies have predominantly used deep learning to analyze EMG signals during the resting state or gestures unrelated to neuromuscular disease diagnosis [28–32]. To the best of our knowledge, few studies have used deep learning to analyze MUAP signals evoked during volitional states, which are more integral for the diagnosis of neuropathy and myopathy.

To overcome the diagnostic limitations of EMG and investigate the feasibility of deep learning in reading of EMG, we developed a deep learning model that could classify individuals into myopathy, neuropathy, or normal state categories based on volitional state EMG signals and compared the classification results of the deep learning model with the results of electrodiagnosis by six physicians.

## Materials and Methods

### Study Design and Preparation

We retrospectively reviewed the electronic medical records of individuals who visited Seoul National University Hospital and underwent EMG between June 2015 and July 2020. Twenty subjects who underwent EMG and were diagnosed with myopathy during this period were selected. Among individuals who were diagnosed with neuropathy and normal, 20 with variable types of neuropathy were selected and 20 were selected randomly for the normal group to match the number of subjects in the myopathy group.

EMG evaluation was performed using the Nicolet EDX EMG system and monopolar needle electrode (Natus, Middleton, WI, USA). The filter was set at 20 Hz (low cut) and 10 kHz (high cut). The EMG signals were recorded with a sampling rate of 48 kHz.

Certified physicians reviewed the EMG data and confirmed the diagnosis for all subjects, and results were used for ground-truth in machine learning. Among the waveform data, electromyographic artifacts that resulted from the needle electrode or patient movements were removed at the beginning and end of EMG data recording. The muscles close to the trunk were labeled as proximal muscles and those far from the trunk were labeled as distal muscles using the elbow and knee joints as reference points of the upper and lower extremities, respectively.

### Ethical Approval, and Informed Consent

This study was approved by the Institutional Review Board of Seoul National University Hospital (No. 2008-055-1147) and was conducted in accordance with the Declaration of Helsinki. The requirement for informed consent was waived owing to the retrospective nature of this study, and private subject information was anonymized before analysis.

### Classification by the Deep Learning Model

The EMG signals were down-sampled to 10 kHz to reduce computational complexity. Data were sliced into segments with lengths of 0.4 s and hop sizes of 0.1 s.

A one-dimensional convolutional neural network (CNN) was used as the deep learning model [33]. The CNN was designed following the basic structures of residual neural and visual geometry group networks, which were verified for the classification of complex data [34, 35]. Our CNN comprised seven spatial reduction blocks, five residual blocks, and fully connected layers (S1 Figure). The Softmax function was applied to the final three-output layer. Hyper-parameters were determined empirically with a learning rate of 10^−3^, batch size of 32, and epochs of 100.

The deep learning model classified subjects through two steps. In the first step, it received multiple segments sliced from the EMG data of individual muscles and annotated the muscle as myopathy, neuropathy, or normal. In the second step, after considering the probabilities of all muscles from individual subjects, a feature vector was produced for classifying the subject into myopathy, neuropathy, or normal. Two methods were used to generate feature vectors, one with and without consideration of muscle location information. The first was to generate a three-dimensional vector (i.e., myopathy/neuropathy/normal) by averaging the probability without muscle location information, and the other was to generate a six-dimensional vector (i.e., myopathy/proximal neuropathy/normal probabilities of proximal muscles and myopathy/neuropathy/normal probabilities of distal muscles) by averaging the probability with muscle location information. A mean probability of 1/3 was imputed to prevent undesired bias for subjects who did not have any muscle probability for some muscle location labels.

### Classification by Physicians

A web-based EMG signal labeling platform that replayed EMG signal was developed to allow physicians to perform electrodiagnosis. Two neurology and four rehabilitation medicine physicians with more than one year of experience in performing and interpreting EMG initially annotated EMG signal of individual muscle and subsequently classified subjects based on the overall muscle annotation results without any clinical information except the EMG signals.

### Performance Assessments

Performance of the deep learning algorithm was compared with results obtained by physicians using the following metrics: accuracy, positive predictive value (precision), sensitivity (recall), specificity, and F1 score. The performance metrics were calculated using the following formulae:

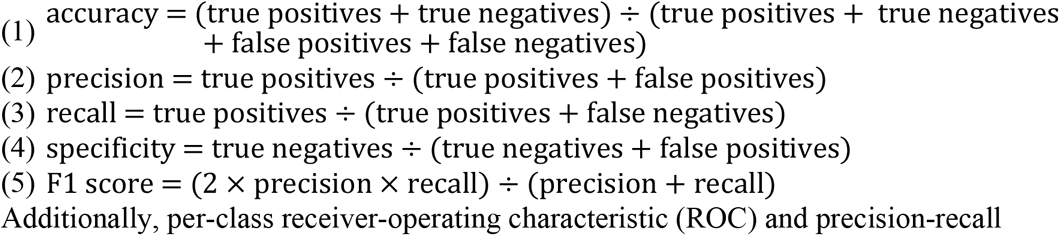

Additionally, per-class receiver-operating characteristic (ROC) and precision-recall curves of the deep learning model were depicted, and the classification results by physicians were also depicted on plots of the deep learning model. Moreover, we compared the results classified by the two versions of the deep learning model with and without muscle location information.

### Statistical Analysis

Statistical analyses were performed using R statistical software (version 4.1.0; R Foundation for Statistical Computing, Vienna, Austria) and Python programming language (version 3.6; Python Software Foundation, Delaware, United States). Normal distribution for the continuous variables was assessed using the Shapiro–Wilk test. Differences in categorical and continuous variables across myopathy, neuropathy, and normal states were assessed using Pearson’s χ^2^ and Kruskal–Wallis tests, respectively. Data are expressed as the mean±standard deviation for continuous variables and as a number (%) for categorical variables. All metrics, except accuracy, had binary classifications and were measured by averaging each class metric using the one-versus-rest method. A p-value < 0.05 was considered statistically significant.

## Results

### Subjects’ Characteristics

Two subjects with neuropathy (n=1) and myopathy (n=1) were excluded as their EMG data was unsuitable for analysis. Overall, 20 normal subjects, 19 subjects with neuropathy, and 19 subjects with myopathy were included. Detailed demographic characteristics of subjects and detailed information of EMG data are presented in Table 1 and S1 Table, respectively.

**Table 1.**
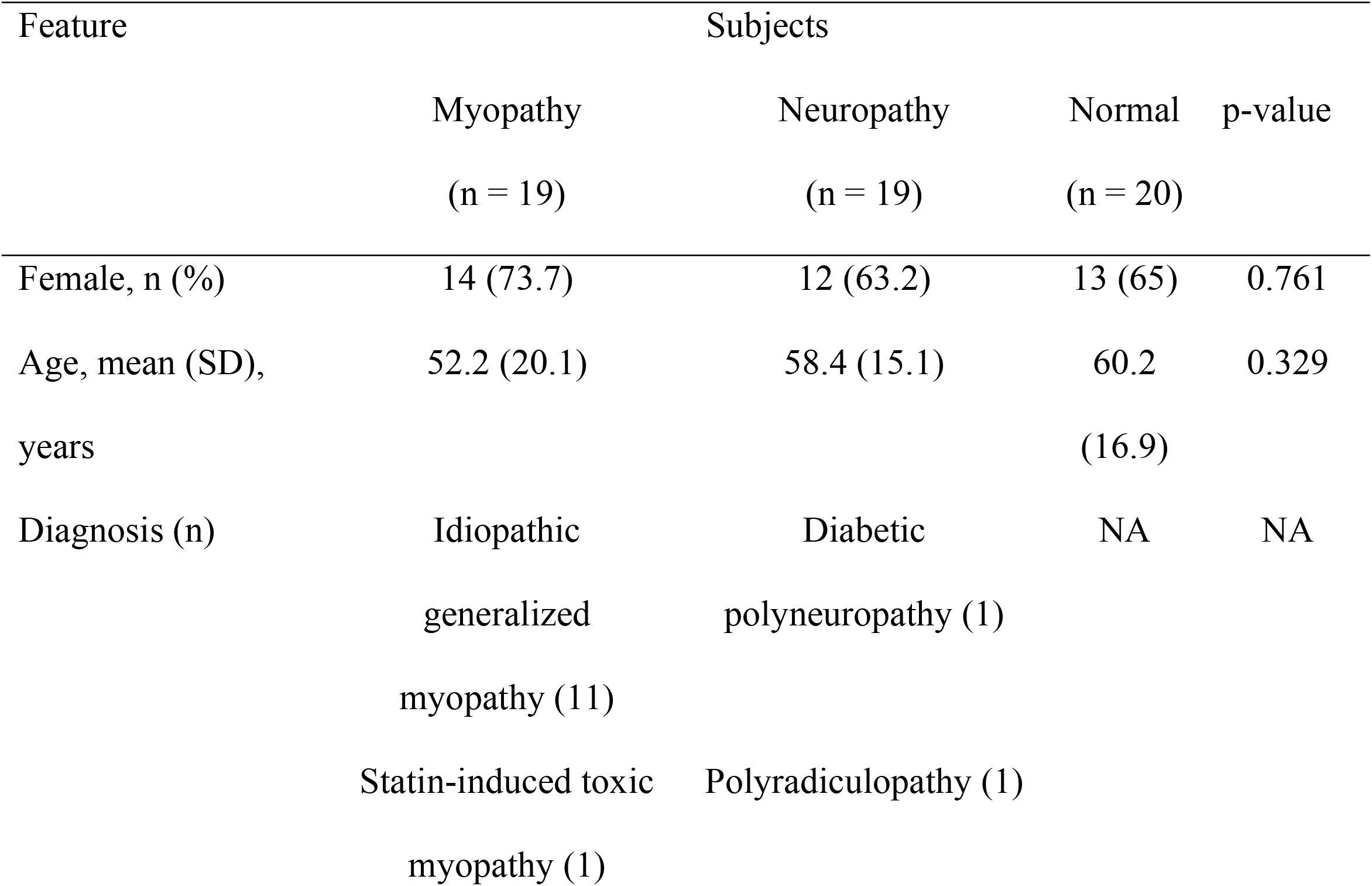
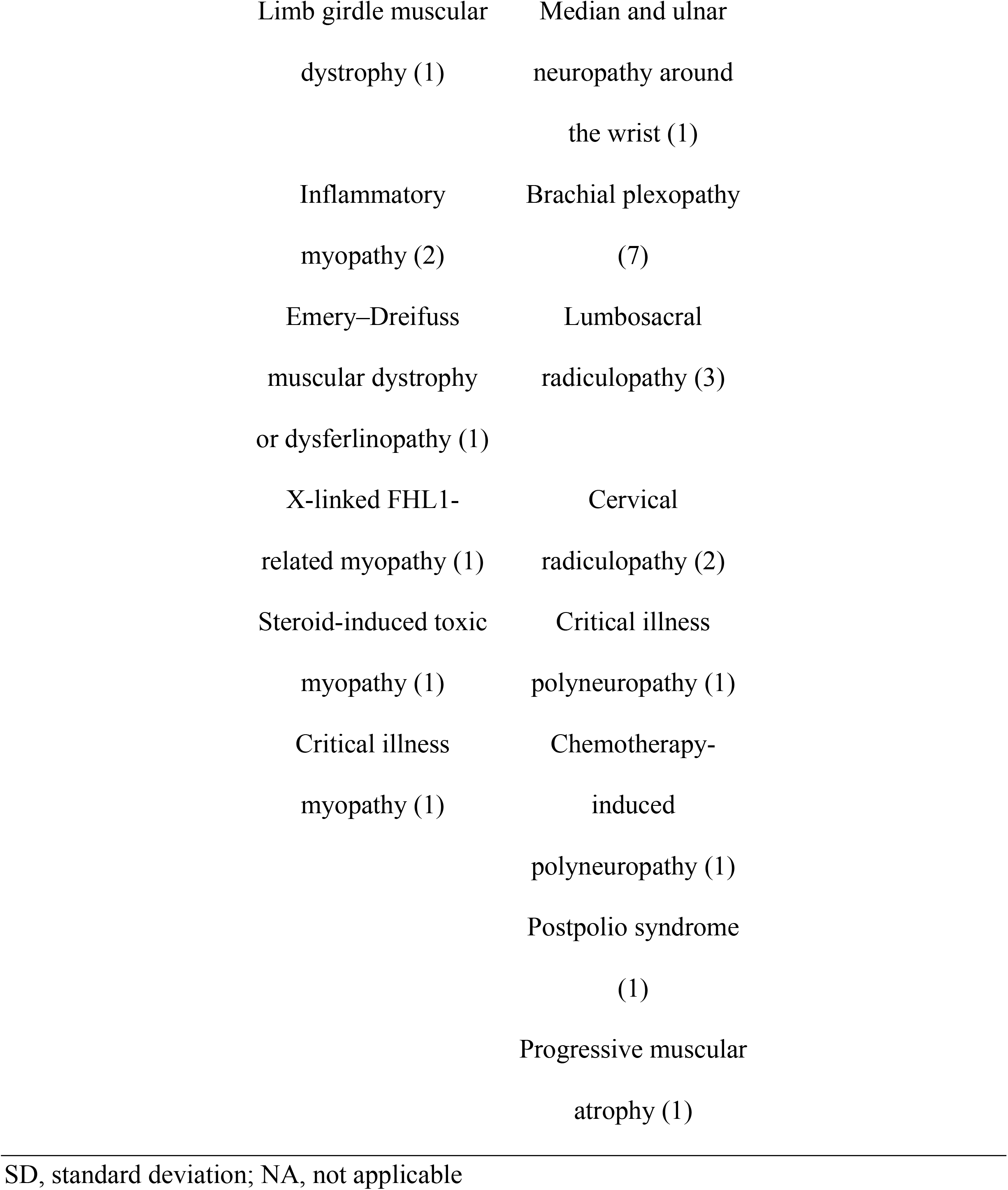
Demographic characteristics of subjects

### Classification Results of the Deep Learning Model without Muscle Location Information

The classification results of the deep learning model that did not consider muscle location information are presented as the following metrics: the accuracy, precision, recall, specificity, and F1 score of the deep learning model were 0.875 (95% confidence interval [CI], 0.864–0.887), 0.820 (95% CI, 0.802–0.839), 0.820 (95% CI, 0.801–0.839), 0.904 (95% CI, 0.898–0.911), and 0.820 (95% CI, 0.801–0.839), respectively (Table 2). The area under the ROC curves of the per-class deep learning model classification results were 0.874 (95% CI, 0.858–0.889), 0.781 (95% CI, 0.723–0.839), and 0.906 (95% CI, 0.899–0.913) for myopathy, neuropathy, and normal, respectively (Fig 1).

**Table 2.** Results of classification results obtained using the deep learning model and by six physicians

|  | Deep learning model<br>(95% CI)<sup>a</sup> | Physicians<br>(95% CI)<sup>b</sup> |
| --- | --- | --- |
| Accuracy | 0.875 (0.864–0.887) | 0.694 (0.664–0.724) |
| Positive predictive value (precision) | 0.820 (0.802–0.839) | 0.602 (0.553–0.651) |
| Sensitivity (recall) | 0.820 (0.801–0.839) | 0.537 (0.491–0.583) |
| Specificity | 0.904 (0.898–0.911) | 0.773 (0.750–0.795) |
| F1 score | 0.820 (0.801–0.839) | 0.524 (0.469–0.578) |

The metrics, except for accuracy, are the averages of the binary classification scores for each class from the one-versus-rest method.
<sup>a</sup>Average results of classification using deep learning model
<sup>b</sup>Average results of classification by six physicians
Abbreviation: CI, confidence interval

**Fig 1.**
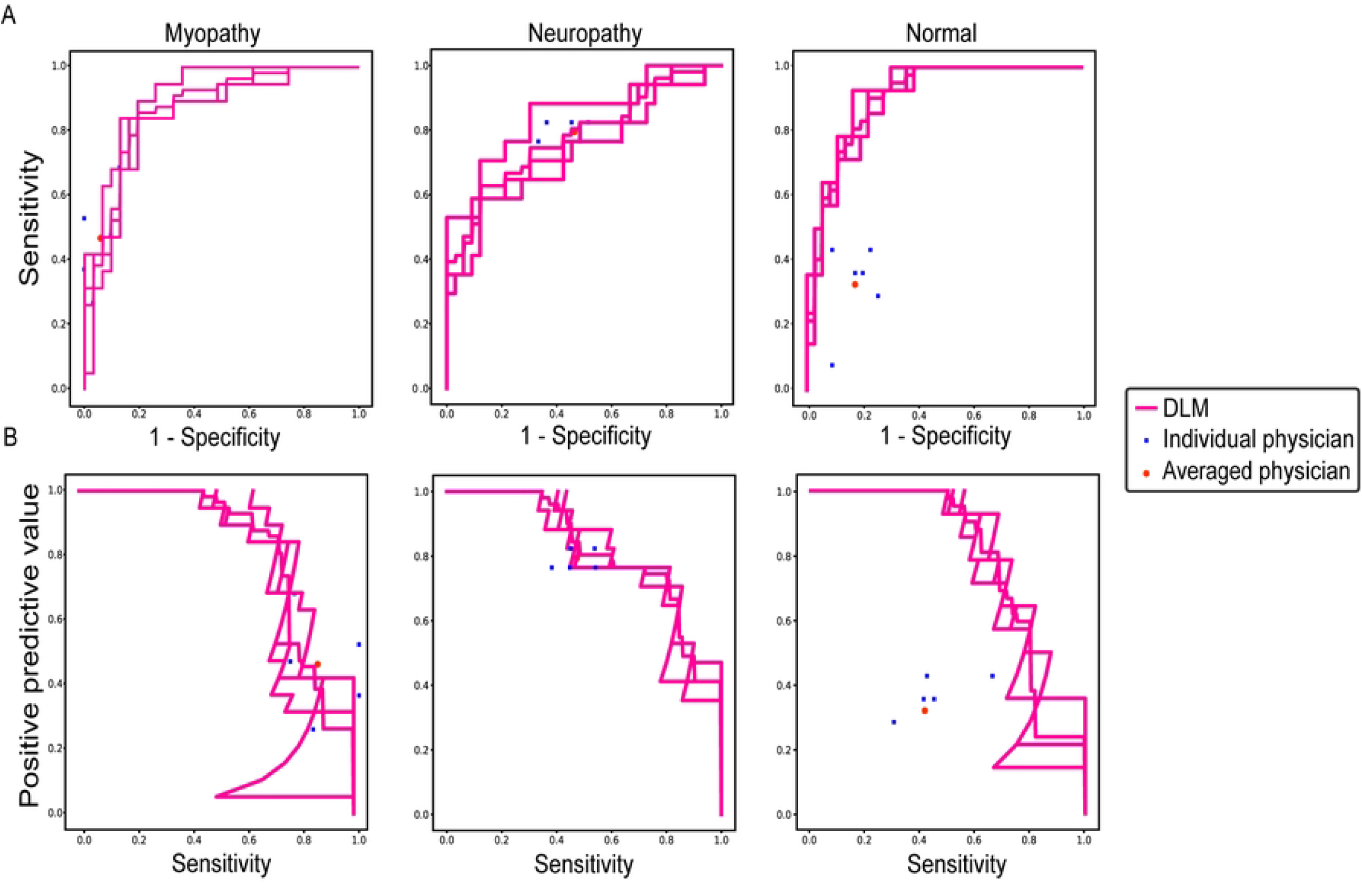
Per-class receiver-operating characteristic and precision-recall curves of the deep learning model and six physicians. (A) Areas under the receiver-operating characteristic curve and (B) precision-recall curve were measured and depicted by dividing all data into myopathy, neuropathy, and normal classes. Individual physician performance is indicated by the blue cross, and average physician performance is indicated by the red dot.Abbreviation: DLM, deep learning model

The overall prediction pattern was identified from the confusion matrix of the deep learning model. The correctly predicted ratios for myopathy, neuropathy, and normal of physicians vs. the deep learning model were 49.49%±13.04% vs. 80.70%±4.96%, 79.41%±2.94% vs. 64.71%±4.80%, and 32.14%±12.20% vs. 69.05%±12.14%, respectively (Fig 2).

**Fig 2.**
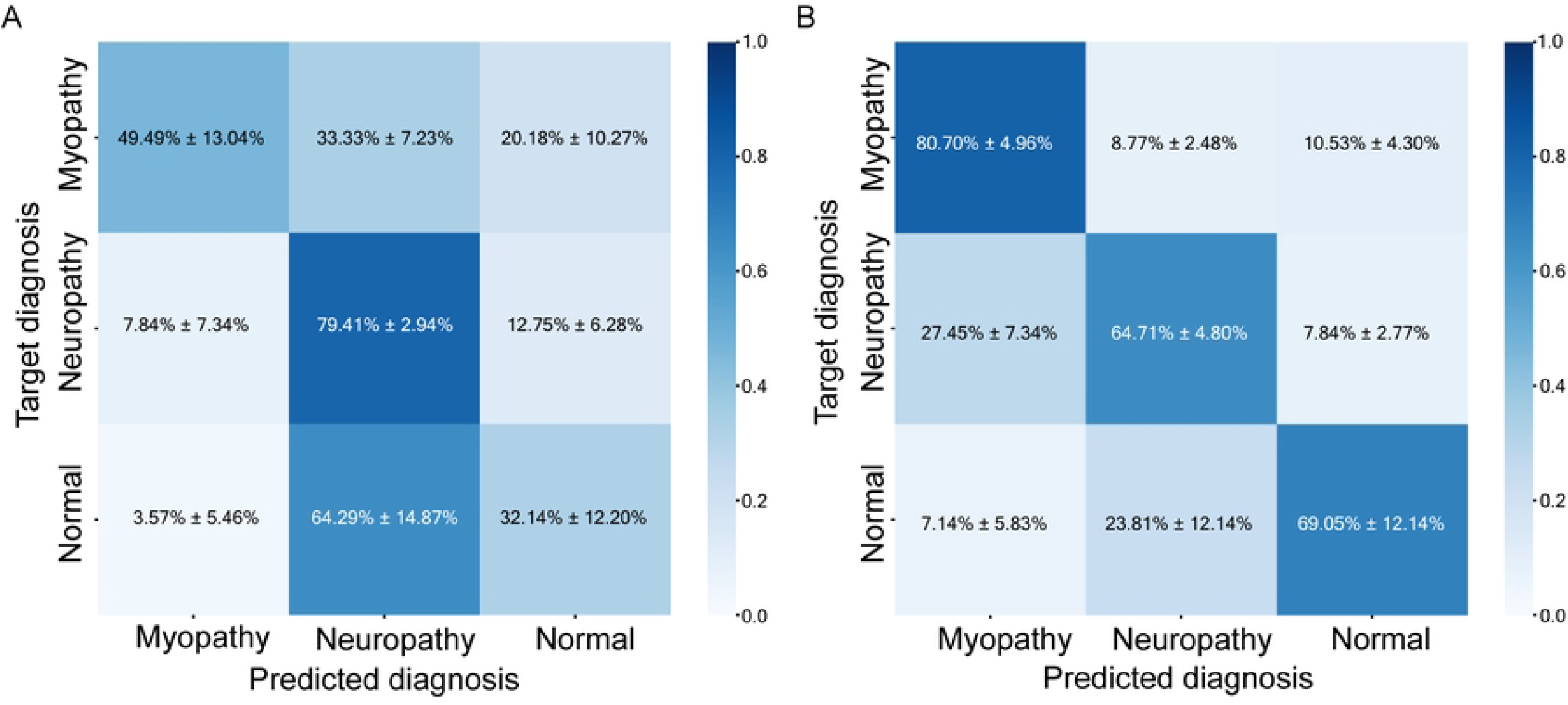
Confusion matrices of the results of physicians and the deep learning model. (A) prediction result of physicians (B) prediction result of the deep learning model

### Classification Results by Physicians

The classification results by physicians are presented as the following metrics: the accuracy, precision, recall, specificity, and F1 score were 0.694 (95% CI, 0.664–0.724), 0.602 (95% CI, 0.553–0.651), 0.537 (95% CI, 0.491–0.583), 0.773 (95% CI, 0.750–0.795), and 0.524 (95% CI, 0.469–0.578), respectively (Table 2). The overall prediction pattern was identified from the confusion matrix of physicians (Fig 2). The correctly predicted ratios for myopathy, neuropathy, and normal by the physicians were 49.49%±13.04%, 79.41%±2.94%, and 32.14%±12.20%, respectively (Fig 2).

### Classification Results of the Deep Learning Model Considering Muscle Location Information

The performance of the deep learning model did not change significantly with the addition of muscle location information. The precision, recall, specificity, and F1 score are shown in S2 Table. The ROC and precision-recall curves are depicted in S2 Figure. The classification results of the deep learning model with muscle location information for myopathy, neuropathy, and normal were not satisfactory (S3 Fig).

### Learned Features of the Deep Learning Model

Waveform characteristics were identified by learned features of the deep learning model. The generated signals were similar to the typical characteristics of neuropathy, myopathy, and normal. Waveforms that were most likely to predict myopathy showed small amplitudes and short durations (Fig 3A), whereas those that most accurately predicted neuropathy showed high amplitudes and long durations (Fig 3B). Thus, we validated the hypothesis that the deep learning model made predictions based on relevant features rather than artifacts. We reviewed misclassified EMG signals to analyze the reasons for the misclassification (S4 Fig).

**Fig 3.**
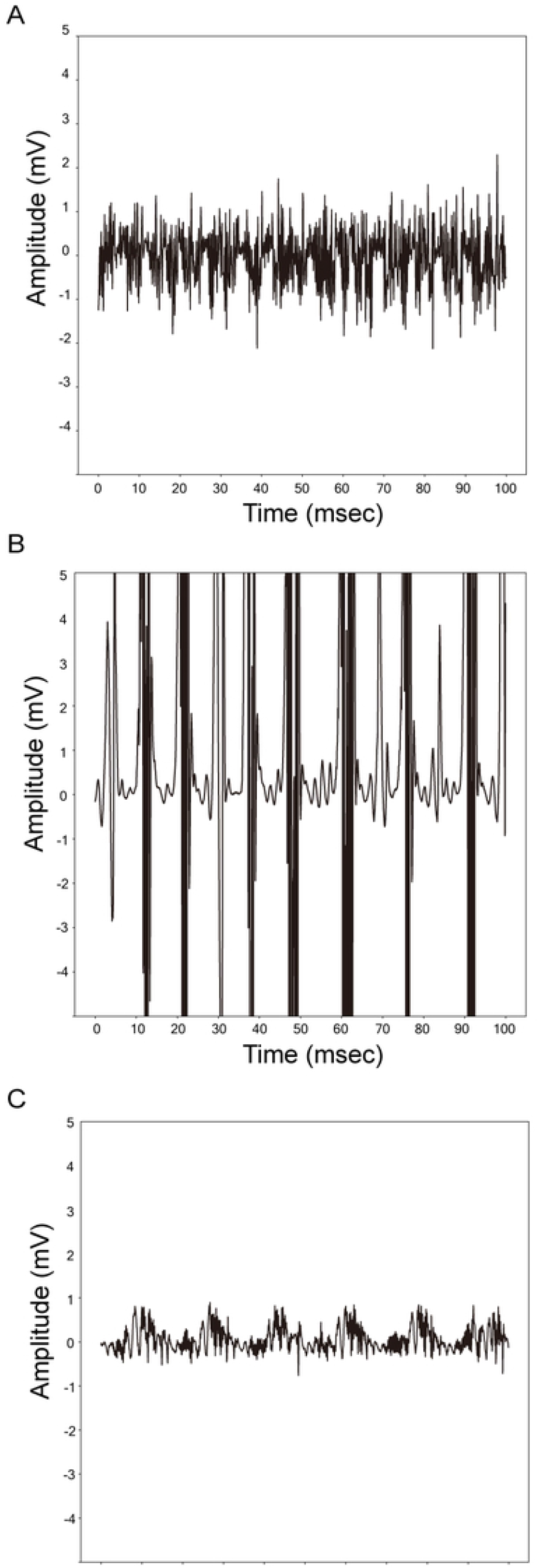
Feature visualization results of the deep learning model. (A) Myopathy (B) Neuropathy (C) Normal.

## Discussion

Overall, the deep learning model outperformed physicians on all performance metrics, with an accuracy of 0.875 (95% CI, 0.864–0.887), which was higher compared to 0.694 (95% CI, 0.664–0.724) for physicians for the classification of EMG signal without additional clinical information.

Deep learning reportedly performs well when used for EMG signal analysis [28–32].

However, previous studies on EMG mainly focused on EMG signals in the resting state [30,31]. We analyzed EMG signals using the deep learning model and confirmed that the model outperformed physicians in diagnosing myopathy and neuropathy.

In some instances, abnormal EMG signals are obtained in limited muscles, and the number of muscles examined may differ among patients. Considering those variations, the EMG results of all tested muscles should be taken into account to diagnose patients with neuromuscular disease. We addressed this variation issue by constructing feature vectors for subjects from the muscle signal prediction probabilities and utilizing an additional classifier to determine the classification results. This method allows the deep learning model to consider all the signals measured from different muscles of different subjects in a consistent format and classify EMG signals based on signal characteristics rather than the number of certain muscle types (i.e., the deep learning model correctly classified whether muscles innervated by specific nerve or spinal roots were more examined or not).

Muscle involvement usually follows typical patterns (i.e., patients with peripheral neuropathy and myopathy predominantly exhibit abnormalities in distal and proximal muscles, respectively) [36]. Although muscle location is meaningful in differentiating between neuropathy and myopathy, the addition of muscle location information did not significantly enhance the performance of the deep learning model. This may be due to the following two reasons. First, some types of myopathies may present with abnormalities of both proximal and distal muscles. For instance, in muscular dystrophy, the distribution of the affected muscles depends on the disease process. Both the proximal and distal muscles are affected in statin-induced and critical illness myopathies. Myotonic dystrophy types 1 and 2 or distal myopathy may affect distal muscles more frequently than proximal muscles [36–38]. Second, the relatively small sample size was insufficient to generate significant features derived from muscle location information.

Several instances where the signals were misclassified by the deep learning model were inspected (S4 Fig). Signals that contained parts with high and low amplitudes were misclassified as neuropathy (S4A Fig) and myopathy (S4B Fig), respectively. The amplitudes of mispredicted EMG signals may dominate the recruitment and interference patterns, thereby resulting in incorrect predictions.

Interestingly, the diagnostic accuracy of the physicians was lower than expected. This can be attributed to the following. First, 38 of the 58 subjects had neuromuscular diseases, which was considerably higher than the real-world prevalence of approximately 200 per 100,000 individuals [39]. Second, electrodiagnosis by physicians differed from the real-world diagnostic process. Physicians usually consider both the EMG signals and additional patient information such as demographics or symptoms, which were absent in this study.

The present study has some limitations. First, we used retrospective data from a single institution. Additional data from other institutions can be used to perform external validation to further verify the model performance. Second, a larger amount of EMG data needs to be examined to demonstrate the stability of performance of the deep learning model in EMG classification. The sample size was insufficient to demonstrate the utility of this deep learning model, and a larger cohort may elicit useful muscle location information. Third, diagnoses were only divided into neuropathy, myopathy, and normal. There are diverse subtypes of neuromuscular diseases, such as chronic inclusion body myositis, ongoing-state dermatomyositis, and late-stage muscular dystrophy, which co-exhibit the MUAPs of myopathy and neuropathy, with short and long durations. Additional EMG data for more specific neuromuscular diseases can improve the performance of deep learning for assisting physicians. Finally, resting-state EMG data should be considered together for more accurate diagnosis. In some neuromuscular diseases, such as Pompe disease, EMG abnormalities may be revealed only in the resting state of the paraspinal muscles rather than that of the limb muscles [36,40]. Future prospective studies with resting and volitional state EMG data can further enhance the applicability of deep learning for EMG electrodiagnosis.

We demonstrated that deep learning could analyze EMG signals in a short time and with high accuracy and that our relatively simple model has the potential to be embedded in an EMG device. Embedding a fast, accurate, and simple deep learning model into an EMG device can allow for clinical assistance to be contained within the device without the need for sharing personal medical information. This would reduce the burden on physicians and lead to a widely applicable, low-cost clinical decision-aiding system for use in medical institutions with limited resources.

## Conclusion

Deep learning has considerable potential to contribute to the development of automatic computer-aided diagnosis systems for patients with neuromuscular diseases.

## Data Availability

All relevant data are within the manuscript and its Supporting Information files.

## Acknowledgements

This work was supported by the Technology Innovation Program (grant number 20016225) and Korea Medical Device Development Fund grant (number 202011B23) by the Ministry of Trade, Industry and Energy (Korea) and the Korean government (the Ministry of Science and ICT; the Ministry of Trade, Industry, and Energy; the Ministry of Health & Welfare; the Ministry of Food and Drug Safety). The study sponsors had no role in the study design, collection, analysis, and interpretation of data, in the writing of the manuscript, and in the decision to submit the manuscript for publication.

## Supporting information

**S1 Table.Characteristics of electromyography data.**

**S2 Table.The results of classification using the deep learning model with and without muscle location information.**

**S1 Fig.Structure of the deep learning model.**

The spatial reduction blocks consisted of convolutional layers, batch normalization, rectified linear unit (ReLU), and max pooling. The residual blocks contained similar layers with added residual connections. The fully connected layers consisted of 512, 256, 64, and 16 sequentially decreasing hidden layer neurons with a leaky ReLU activation function. Early stopping was performed by evaluating the accuracy of the validation set after every 30 updates, and the patience value was set to 100. Cross-entropy loss was used as the loss function, with class weights applied in an inversely proportional manner to the number of signal segments from the training set. Deep learning performance was measured through 5 × 3-fold cross validation due to the small number of subjects

**S2 Fig.Per-class receiver operating characteristic (ROC) and precision-recall curves of the deep learning model with and without muscle location information.**

The areas under the (A) ROC and (B) precision-recall curves were measured and depicted by dividing all data into the myopathy, neuropathy, and normal groups.

Individual physician performance is indicated by the blue cross, and average physician performance is indicated by the red dot.

ROC and precision-recall curves of the deep learning model depending on whether muscle location information was considered (green lines, with muscle location) or not (pink lines, without muscle location).

Abbreviations: DLM, deep learning model; ML, muscle location

**S3 Fig.Confusion matrices of the deep learning model (A) without and (B) with muscle location information.**

**S4 Fig.Examples of electromyographic signals mispredicted by the deep learning model.**

Electromyographic signals mispredicted as (A) and (B) neuropathy, (C) and (D) myopathy, and (E) and (F) normal signals.

## References

1. Daube JR, Rubin DI. Needle electromyography. Muscle Nerve. 2009;39: 244–270. doi: 10.1002/mus.21180.

2. Kimura J. Electrodiagnosis in diseases of nerve and muscle: principles and practice. New York: Oxford University Press; 2013.

3. Mills KR. The basics of electromyography. J Neurol Neurosurg Psychiatry. 2005;76 Suppl 2: ii32–ii35. doi: 10.1136/jnnp.2005.069211.

4. Oh SJ. Clinical electromyography: nerve conduction studies. Philadelphia: Lippincott Williams & Wilkins; 2003.

5. Rubin DI. Needle electromyography: basic concepts. Handb Clin Neurol. 2019;160: 243–256. doi: 10.1016/B978-0-444-64032-1.00016-3.

6. Whittaker RG. The fundamentals of electromyography. Pract Neurol. 2012;12: 187–194. doi: 10.1136/practneurol-2011-000198.

7. Aminoff MJ, Goodin DS, Parry GJ, Barbaro NM, Weinstein PR, Rosenblum ML. Electrophysiologic evaluation of lumbosacral radiculopathies: electromyography, late responses, and somatosensory evoked potentials. Neurology. 1985;35: 1514–1518. doi: 10.1212/wnl.35.10.1514.

8. Bromberg MB. The motor unit and quantitative electromyography. Muscle Nerve. 2020;61: 131–142. doi: 10.1002/mus.26718.

9. Gutiérrez-Gutiérrez G, Barbosa López C, Navacerrada F, Miralles Martínez A. Use of electromyography in the diagnosis of inflammatory myopathies. Reumatol Clin. 2012;8: 195–200. doi: 10.1016/j.reuma.2011.10.012.

10. Leblhuber F, Reisecker F, Boehm-Jurkovic H, Witzmann A, Deisenhammer E. Diagnostic value of different electrophysiologic tests in cervical disk prolapse. Neurology. 1988;38: 1879–1881. doi: 10.1212/wnl.38.12.1879.

11. Sawada K, Horii M, Imoto D, Ozaki K, Toyama S, Saitoh E, et al. Usefulness of electromyography to predict future muscle weakness in clinically unaffected muscles of polio survivors. PM R. 2020;12: 692–698. doi: 10.1002/pmrj.12281.

12. Tonzola RF, Ackil AA, Shahani BT, Young RR. Usefulness of electrophysiological studies in the diagnosis of lumbosacral root disease. Ann Neurol. 1981;9: 305–308. doi: 10.1002/ana.410090317.

13. Haig AJ, Tong HC, Yamakawa KS, Quint DJ, Hoff JT, Chiodo A, et al. The sensitivity and specificity of electrodiagnostic testing for the clinical syndrome of lumbar spinal stenosis. ySpine (Phila Pa 1976). 2005;30:2667–2676. doi: 10.1097/01.brs.0000188400.11490.5f.

14. Kendall R, Werner RA. Interrater reliability of the needle examination in lumbosacral radiculopathy. Muscle Nerve. 2006;34: 238–241. doi: 10.1002/mus.20554.

15. Nirkko AC, Rösler KM, Hess CW. Sensitivity and specificity of needle electromyography: a prospective study comparing automated interference pattern analysis with single motor unit potential analysis. Electroencephalogr Clin Neurophysiol. 1995:97: 1–10. doi: 10.1016/0924-980x(94)00248-6.

16. Arthur KC, Calvo A, Price TR, Geiger JT, Chiò A, Traynor BJ. Projected increase in amyotrophic lateral sclerosis from 2015 to 2040. Nat Commun. 2016;7: 12408. doi: 10.1038/ncomms12408.

17. Longinetti E, Fang F. Epidemiology of amyotrophic lateral sclerosis: an update of recent literature. Curr Opin Neurol. 2019;32: 771–776. doi: 10.1097/WCO.0000000000000730.

18. Parker MJS, Oldroyd A, Roberts ME, Ollier WE, New RP, Cooper RG, et al. Increasing incidence of adult idiopathic inflammatory myopathies in the City of Salford, UK: a 10-year epidemiological study. Rheumatol Adv Pract. 2018;2: rky035. doi: 10.1093/rap/rky035.

19. Rose L, McKim D, Leasa D, Nonoyama M, Tandon A, Bai YQ, et al. Trends in incidence, prevalence, and mortality of neuromuscular disease in Ontario, Canada: a population-based retrospective cohort study (2003-2014). PLoS One. 2019;14: e0210574. doi: 10.1371/journal.pone.0210574.

20. Alfaras M, Soriano MC, Ortí n S. A fast machine learning model for ECG-based heartbeat classification and arrhythmia detection. Front Phys. 2019;7: 103. doi: 10.3389/fphy.2019.00103.

21. Lu X, Wu Y, Yan R, Cao S, Wang K, Mou S. et al. Pulse waveform analysis for pregnancy diagnosis based on machine learning. 2018 IEEE 3rd Advanced Information Technology, Electronic and Automation Control Conference (IAEAC). 2018; 1075–1079.

22. Gemein LAW, Schirrmeister RT, Chrabąszcz P, Wilson D, Boedecker J, Schulze-Bonhage A, et al. Machine-learning-based diagnostics of EEG pathology. Neuroimage. 2020;220: 117021. doi: 10.1016/j.neuroimage.2020.117021.

23. Roy Y, Banville H, Albuquerque I, Gramfort A, Falk TH, Faubert J. Deep learning-based electroencephalography analysis: a systematic review. J Neural Eng. 2019;16: 051001. doi: 10.1088/1741-2552/ab260c.

24. Bien N, Rajpurkar P, Ball RL, Irvin J, Park A, Jones E, et al. Deep-learning-assisted diagnosis for knee magnetic resonance imaging: development and retrospective validation of MRNet. PLoS Med. 2018;15: e1002699. doi: 10.1371/journal.pmed.1002699.

25. Hannun AY, Rajpurkar P, Haghpanahi M, Tison GH, Bourn C, Turakhia MP, et al. Cardiologist-level arrhythmia detection and classification in ambulatory electrocardiograms using a deep neural network. Nat Med. 2019;25: 65–59. doi: 10.1038/s41591-018-0268-3.

26. Rajpurkar P, Irvin J, Ball RL, Zhu K, Yang B, Mehta H, et al. Deep learning for chest radiograph diagnosis: a retrospective comparison of the CheXNeXt algorithm to practicing radiologists. PLoS Med. 2018;15: e1002686. doi: 10.1371/journal.pmed.1002686.

27. Ribeiro AH, Ribeiro MH, Paixão GMM, Oliveira DM, Gomes PR, Canazart JA, et al. Automatic diagnosis of the 12-lead ECG using a deep neural network. Nat Commun. 2020;11: 1760. doi: 10.1038/s41467-020-15432-4.

28. Akef Khowailed I, Abotabl A. Neural muscle activation detection: a deep learning approach using surface electromyography. J Biomech. 2019;95: 109322. doi: 10.1016/j.jbiomech.2019.109322.

29. Atzori M, Cognolato M, Müller H. Deep learning with convolutional neural networks applied to electromyography data: a resource for the classification of movements for prosthetic hands. Front Neurorobot. 2016;10: 9. doi: 10.3389/fnbot.2016.00009.

30. Nam S, Sohn MK, Kim HA, Kong HJ, Jung IY. Development of artificial intelligence to support needle electromyography diagnostic analysis. Healthc Inform Res. 2019;25: 131–138. doi: 10.4258/hir.2019.25.2.131.

31. Nodera H, Osaki Y, Yamazaki H, Mori A, Izumi Y, Kaji R. Deep learning for waveform identification of resting needle electromyography signals. Clin Neurophysiol. 2019;130: 617–623. doi: 10.1016/j.clinph.2019.01.024.

32. Wei W, Dai Q, Wong Y, Hu Y, Kankanhalli M, Geng W. Surface-electromyography-based gesture recognition by multi-view deep learning. IEEE Trans Biomed Eng. 2019;66: 2964–2973. doi: 10.1109/TBME.2019.2899222.

33. Krizhevsky A, Sutskever I, Hinton GE. Imagenet classification with deep convolutional neural networks. Commun ACM. 2017;60: 84–90. doi: 10.1145/3065386.

34. He K. Deep residual learning for image recognition. Proceedings of the IEEE Conference on Computer Vision and Pattern Recognition, 2016;770–778.

35. Simonyan K, Zisserman A. Very deep convolutional networks for large-scale image recognition. Arxiv. 2014. doi: 10.48550/arXiv.1409.1556.

36. Paganoni S, Amato A. Electrodiagnostic evaluation of myopathies. Phys Med Rehabil Clin N Am. 2013;24: 193–207. doi: 10.1016/j.pmr.2012.08.017.

37. Logigian EL, Ciafaloni E, Quinn LC, Dilek N, Pandya S, Moxley RT, et al. Severity, type, and distribution of myotonic discharges are different in type 1 and type 2 myotonic dystrophy. Muscle Nerve. 2007;35: 479–485. doi: 10.1002/mus.2072220722.

38. Dimachkie MM, Barohn RJ. Distal myopathies. Neurol Clin. 2014;32: 817–842, x. doi: 10.1016/j.ncl.2014.04.004.

39. Carey IM, Banchoff E, Nirmalananthan N, Harris T, DeWilde S, Chaudhry UAR, et al. Prevalence and incidence of neuromuscular conditions in the UK between 2000 and 2019: a retrospective study using primary care data. PLoS One. 2021;16: e0261983. doi: 10.1371/journal.pone.0261983.

40. Hobson-Webb LD, Dearmey S, Kishnani PS. The clinical and electrodiagnostic characteristics of Pompe disease with post-enzyme replacement therapy findings. Clin Neurophysiol. 2011;122: 2312–2317. doi: 10.1016/j.clinph.2011.04.016.

